# Percutaneous coronary intervention of patients with chronic total occlusion is associated with higher mortality and complications despite propensity score matching

**DOI:** 10.1101/2025.10.05.25337387

**Authors:** Diego Garcia, Dylan Langland, Mehrtash Hashemzadeh, Mohammad Reza Movahed

## Abstract

**Background:** The rate of percutaneous coronary intervention (PCI) of chronic total occlusions (CTO) is increasing, yet its clinical value remains controversial. CTO-PCI is associated with high procedural risk, such as perforation and tamponade. The goal of this study was to compare complications and mortality of patients undergoing CTO-PCI with patients without CTO undergoing PCI (non-CTO-PCI) by using propensity score matching.

**Methods:** The national Inpatient Sample database (NIS), years 2016-2020, was studied using International Classification of Diseases, Tenth Revision Codes. Propensity Score Matching was used to compare patients with CTO-PCI with patients non-CTO-PCI.

**Results:** Among 519,620 PCI hospitalizations, 259,810 had CTO-PCI. After matching, CTO-PCI had higher mortality (OR 1.13; 95% CI 1.05–1.22; p=0.001) and higher odds of myocardial infarction (OR 1.97; 1.64–2.38), perforation (OR 3.36; 2.63–4.28), tamponade (OR 2.07; 1.63– 2.65), bleeding (OR 1.88; 1.64–2.15), and respiratory failure (OR 1.80; 1.45–2.24); all p<0.001. Overall risk doubled for any complication (OR 2.04; 1.85–2.25) and rose to 42% for MACE (OR 1.42; 1.34–1.51). When perforation or tamponade occurred, mortality was striking much higher in CTO-PCI compared to non-CTO-PCI (2.43% vs 0.55% and 22.86% vs 1.17%; both p<0.001).

**Conclusions:** Propensity-matched national cohort confirmed that CTO-PCI was associated with higher in-hospital complications and mortality compared to non-CTO-PCI, mostly driven by perforation and tamponade. These findings support the previous report that CTO-PCI is associated with worse outcomes.

## Introduction

A chronic total occlusion (CTO) of the coronary artery is a severe or complete luminal blockage that prevents meaningful opacification of the distal vessel bed (Thrombolysis in Myocardial Infarction flow grades [TIMI] 0-1) for ≥ 3 months.^1,2^ In studies completed on patients with significant coronary artery disease who undergo percutaneous coronary intervention (PCI), it has been shown that as many as 52% of patients have a CTO.^3^ It is also apparent that the number of patients with CTO who undergo PCI has continued to increase over the years, likely as a result of improvements in catheter technology and revascularization approaches.^4-6^ For example, a study completed in 2013 demonstrated that 30% of patients with CTO had a PCI.^7^

The benefits of successful PCI for those patients with CTO are very limited, if any, including angina relief or improved quality of life.^8-11^ However, the methodology of studies showing such benefits tends to compare outcomes of successful vs unsuccessful PCI for CTO instead of comparing PCI for CTO with optimal medical therapy (OMT) or no therapy. Those studies that do explore PCI for CTO against OMT or no therapy have found no significant difference in hard outcomes like mortality. ^12,13^

Moreover, PCI in the setting of CTO has the potential for many serious negative ramifications that can result from the procedure, such as pseudoaneurysm, retroperitoneal bleed, periprocedural MI, coronary dissection, arrhythmias, stent thrombosis, stroke, and many more.^14-16^ The rate of coronary artery perforation, for example, is higher in PCI in the setting of CTO compared to normal PCI. A retrospective observational cohort study of CTO-PCI interventions revealed higher mortality and complications in comparison to non-CTO PCI despite multivariable adjustment. However, despite being the largest study ever reported, it was not a propensity score match.^17,18^ In addition to the likelihood of severe complications, PCI for patients with CTO has been shown to have a first attempt failure rate as high as 24.9%.^17^ In accordance with conflicting studies that make different recommendations regarding the use of PCI in the setting of CTO, there is no professional consensus, and current recommendations remain vague.^19^ The goal of this study was to perform the largest retrospective data analysis comparing CTO PCI to non-CTO PCI for the occurrence of in-hospital mortality and complications using propensity score matching, adjusting for numerous baseline characteristics and high-risk features.

## Methods

### Data Source

The study cohort was derived from the National Inpatient Sample (NIS), Healthcare Cost and Utilization Project (HCUP), and Agency for Healthcare Research and Quality database (AHRQ). The NIS is the largest U.S. database of hospital stays, containing a de-identified sample of discharge records from hospitals nationwide; it’s one of several files in HCUP, a family of de-identified hospital databases and tools curated from state and hospital data systems; and AHRQ is the federal agency that sponsors HCUP and makes these datasets and methods available. HCUP NIS data are publicly available and deidentified, and thus, the study was exempt from institutional review board approval. The NIS database contains weighted, discharge‐level data approximating ∼35 million hospitalizations annually, derived from a 20% stratified sample of discharges from U.S. community hospitals, and is designed to yield national estimates representative of ∼98% of the U.S. population.

### Study Population

We queried the NIS for the 2016–2020 releases and constructed the cohort using both ICD-10-CM diagnoses and ICD-10-PCS procedure codes. Hospitalizations with PCI were identified by ICD-10-PCS codes 02703(4–7)Z, 02703(D–G)Z, 02703TZ, 02713(4–7)Z, 02713(D–G)Z, 02713TZ, 02723(4–7)Z, 02723(D–G)Z, 02723TZ, 02733(4–7)Z, 02733(D–G)Z, 02733TZ, 02H(0–3)3DZ, 02H(0–3)3YZ, 027(0–3)3ZZ, 02C(0–3)3Z7, 02C(0–3)3ZZ, and 02F(0–3)3ZZ. Cases of CTO were flagged using ICD-10-CM I25.82. Analyses were restricted to patients older than 30 years, and demographic variables extracted for description included age, sex, and race.

### Study Outcomes

We evaluated in-hospital all-cause mortality and procedure-related complications identified by ICD-10-CM codes: postprocedural myocardial infarction (I97.89), contrast-induced nephropathy (N99.0), cardiac perforation (I97.51), procedural bleeding (I97.410, I97.411, I97.610, I97.611, I97.630, I97.631), cardiac tamponade (I31.4), acute postprocedural respiratory failure (J95.821), and postprocedural ischemic stroke (I97.821). We defined major adverse cardiac events (MACE) as a composite of all measured cardiac complications together with in-hospital mortality. For propensity score matching, the mortality analysis was adjusted using baseline characteristics and established high-risk features, including age, sex, race, diabetes mellitus, chronic kidney disease, systolic heart failure, three-vessel PCI, hypertension, chronic obstructive pulmonary disease, ST-elevation myocardial infarction, non–ST-elevation myocardial infarction, prior PCI, prior coronary artery bypass grafting, anemia, smoking status, atrial fibrillation/flutter, valvular heart disease, endocarditis, and history of stroke.

## Results

Among 519,620 patients undergoing PCI between 2016 and 2020, 259,810 underwent intervention for chronic total occlusion (CTO). After propensity score matching to balance baseline characteristics and high-risk factors such as diabetes, chronic kidney disease, systolic heart failure, 3-vessel PCI, hypertension, chronic obstructive pulmonary disease, ST-elevation myocardial infarction, non-ST-elevation myocardial infarction, prior PCI, history of coronary artery bypass graft, history of anemia, smoking status, atrial fibrillation/flutter, valvular heart disease, endocarditis, history of stroke, and other cardiovascular risk factors, CTO intervention remained independently associated with all-cause mortality (OR 1.13; 95% CI, 1.05–1.22; p = 0.001, Table 1). Furthermore, after propensity score matching, CTO PCI also carried increased odds of major postprocedural complications, including myocardial infarction (OR 1.97, 95% CI 1.64–2.38), perforation (OR 3.36, 95% CI 2.63–4.28), tamponade (OR 2.07, 95% CI 1.63–2.65), bleeding (OR 1.88, 95% CI 1.64–2.15), and respiratory failure (OR 1.80, 95% CI 1.45–2.24), all *p* < 0.001. Contrast-induced nephropathy and cerebrovascular infarction were also higher but didn’t reach statistical significance due to rare reporting (OR 1.35, CI 0.8-2.2, p=0.26 and OR 2.4, CI 0.99-5.94, p =0.052). Overall, CTO PCI was associated with a twofold higher risk of any postprocedural complication (OR 2.04, 95% CI 1.85–2.25) and a 42% higher risk of major adverse cardiac events (MACE; OR 1.42, 95% CI 1.34–1.51). The highest mortality in CTO PCI vs non-CTO PCI was related to perforations (2.43% vs 0.55%, p<0.001) and cardiac tamponade (22.86% vs 1.17%, p<0.001, Table 2). Figure summarizing the odds ratio of mortality related to CTP-PCI compared to non-CTO PCI.

**Table 1:** Study Cohort Baseline Characteristics and Propensity Score Matched Risk Factors.

|  | Total<br>(N=519,620) | No CTO<br>(N=259,810) | CTO<br>(N=259,810) | P-value |
| --- | --- | --- | --- | --- |
| Age |  |  |  | <0.001 |
| Mean±SD | 66.68±11.81 | 66.39±11.83 | 66.97±11.76 |  |
| Median (IQR) | 67(59-75) | 66(58-75) | 67(59-76) |  |
| Age Group |  |  |  | <0.001 |
| 30-45 | 4.16% | 4.54% | 3.79% |  |
| 46-60 | 25.86% | 26.16% | 25.56% |  |
| 61+ | 69.97% | 69.30% | 70.64% |  |
| Gender |  |  |  | <0.001 |
| Male | 70.60% | 67.96% | 73.24% |  |
| Female | 29.40% | 32.04% | 26.76% |  |
| Race |  |  |  | <0.001 |
| White | 75.27% | 75.22% | 75.33% |  |
| Black | 10.18% | 10.49% | 9.87% |  |
| Hispanic | 8.02% | 8.13% | 7.90% |  |
| Asian/Pac Isl | 2.64% | 2.39% | 2.88% |  |
| Native American | 0.64% | 0.64% | 0.63% |  |
| Others | 3.26% | 3.13% | 3.38% |  |
| **Propensity Score Matched Risk Factors** |  |  |  |  |
| STEMI | 21.61% | 21.62% | 21.60% | 0.95 |
| Non-STEMI | 23.80% | 23.83% | 23.76% | 0.82 |
| Systolic Heart Failure | 28.86% | 28.87% | 28.86% | 0.97 |
| PCI Three Vessel | 2.30% | 2.21% | 2.40% | 0.06 |
| Diabetes | 47.26% | 47.30% | 47.22% | 0.81 |
| Chronic Kidney Disease | 27.43% | 27.45% | 27.41% | 0.89 |
| Hypertension | 87.24% | 87.29% | 87.20% | 0.69 |
| COPD | 20.44% | 20.41% | 20.47% | 0.81 |
| Prior Percutaneous Coronary Intervention | 50.07% | 50.05% | 50.10% | 0.89 |
| History of Coronary Artery Bypass Graft | 22.80% | 22.77% | 22.84% | 0.79 |
| History of Anemia | 0.02% | 0.02% | 0.02% | 0.65 |
| History of Cardiomyopathy | 0.87% | 0.85% | 0.89% | 0.59 |
| Previous Smokers | 30.39% | 30.37% | 30.40% | 0.93 |
| Current Smokers | 17.26% | 17.22% | 17.30% | 0.73 |
| Atrial Fibrillation/Flutter | 22.97% | 22.97% | 22.97% | 0.99 |
| Valvular Heart Disease | 12.20% | 1.23% | 12.16% | 0.72 |
| Endocarditis | 0.05% | 0.05% | 0.05% | 0.78 |
| History of Stroke | 1.66% | 1.63% | 1.69% | 0.45 |
| Angina | 0.05% | 0.05% | 0.05% | 0.89 |
| Hyperlipidemia | 77.14% | 77.17% | 77.11% | 0.83 |
| Alcohol | 2.87% | 2.87% | 2.86% | 0.88 |

**Table 2:**
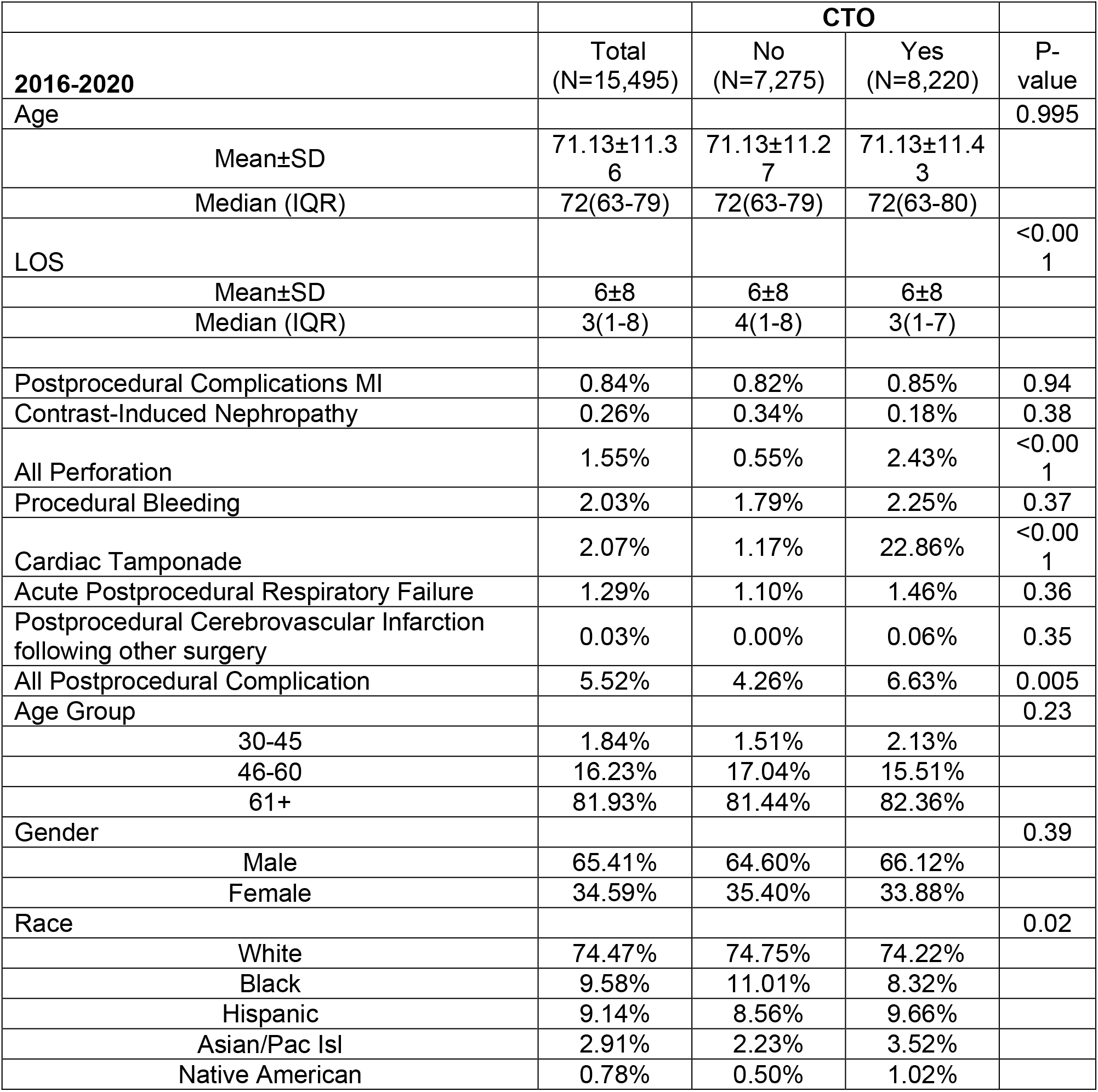
Mortality based on the complications in patients with chronic total occlusion (CTO) percutaneous intervention (PCI) vs no CTO PCI.

## Discussion

Persistent ambiguity in contemporary guidance provides the rationale for this study. On one hand, prior work documents substantial periprocedural hazards—consistent with our observed excess complications with CTO-PCI.^14–16^ On the other hand, several studies (reviewed below) have favored CTO-PCI. This divergence in the evidence base helps explain why the 2021 ACC/AHA/SCAI guideline issued only a weak recommendation and why subsequent commentary has called for greater oversight of indications for CTO-PCI.^19,20^

Simsek et al. systematic review and meta-analysis of clinical outcomes on patients undergoing CTO PCI vs no CTO PCI concludes that there is a need for appropriately powered RCTs to conclusively determine the impact of CTO-PCI on clinical outcomes.^21^ Multiple well-performed trials subsequently built evidence for a lack of significant benefit of PCI: DECISION-CTO showed no difference in major events between CTO-PCI plus medical therapy and medical therapy alone;^22^ EXPLORE showed no improvement in LV function with routine CTO intervention;^23^ REVASC failed to show regional functional recovery;^24^ and the double-blind ORBITA trial found no symptomatic or exercise-time benefit versus a placebo procedure after optimized medical therapy.^25^

This contrasts with studies that have overt design limitations, such as Xenogiannis et al., which compares successful vs unsuccessful CTO-PCI—an approach prone to selection bias.^26^ Juricic’s COMET-CTO, although an RCT, can be misleading in favor of PCI over OMT: it is single-center with a small sample (n=100) and not powered for MACE or all-cause mortality;^27^ exclusions likely selected participants more apt to tolerate PCI, biasing toward PCI over OMT. Park’s single-center, observational follow-up reported late survival benefit with PCI; however, baseline imbalances (fewer prior MI, lower LV dysfunction, younger age, more guideline-directed therapy) and lack of longitudinal therapy data limit interpretation and generalizability.^28^ A reply has argued that, given the lack of mortality benefit and high procedural risk, CTO-PCI should be discouraged, and further critiques highlight possible bias and underreporting of adverse events.^29–33^

We then turn to larger observational data comparing strategies. Nathan et al. reported the largest retrospective analysis comparing CTO vs non-CTO PCI, demonstrating higher mortality and multiple complications with CTO-PCI, mirroring concerns raised by guidelines and trials above.^29^ However, their study was not propensity-matched. Our propensity-matched study not only confirmed their finding but also found a higher independent risk for mortality.

One of the very striking findings in our results, not previously reported, is the fact that Perforations and cardiac tamponade occurring in the setting of CTO PCI were much more deadly than in the setting of non-CTO-PCI. For example, cardiac tamponade in the setting of CTO-PCI had a 22.86% mortality rate in comparison to non-CTO-PCI, with a mortality rate of only 1.17% (Table 2). This suggests that the degree of perforations in the setting of CTO-PCI is much higher than in non-CTO-PCI, leading to a higher death rate. In CTO interventions, coronary perforations are disproportionately high-grade by the Ellis classification—Grade I (extraluminal crater without extravasation), Grade II (pericardial/myocardial blush), Grade III with frank contrast streaming, and Grade III-CS with cavity spilling—because wire-escalation with stiff/hydrophilic wires, subintimal tracking/re-entry, retrograde techniques, and long, calcified lesions increase the risk of large-caliber vessel injury.^34^ High-grade Ellis III/III-CS events are most strongly linked to hemodynamic collapse, severe tamponade, need for pericardiocentesis or emergency surgery, and death—mechanistically explaining the higher mortality observed with CTO-PCI.^34^ In contemporary CTO registries, perforation occurs in ∼4.9% of cases; ∼50% involve the target CTO vessel, nearly 50% relate to the retrograde approach, guidewire exit is the most common mechanism, >50% require active management (prolonged balloon inflation or covered stents), tamponade needing pericardiocentesis occurs in ∼14%, and in-hospital mortality is ∼4%.^35, 36^ Therefore, it is evident that there are significant complications associated with PCI in the setting of CTO. OMT, on the other hand, has been frequently used due to its proven efficacy, and when feasible, and patient safety.^29^ Taken together—and reinforced by our propensity-matched national analysis—CTO-PCI does not confer a consistent survival advantage, while the elevated risks appear not to be simply due to sicker baseline characteristics. Finally, risk-stratification tools such as the Antegrade CTO Score provide a structured means of predicting procedural success or failure; incorporating such scores highlights that patient selection and technical difficulty remain central determinants of CTO-PCI outcomes, beyond the broader morbidity and mortality trends demonstrated in our analysis.^2^ Clinical practice should therefore remain cautious, reserving CTO-PCI for highly selected patients.

## Limitations

Several limitations of this study must be acknowledged, particularly those related to the HCUP NIS database. Importantly, the database captures only patients who have been admitted. While PCI for chronic total occlusion typically requires hospitalization, other PCIs are often performed in the outpatient setting and thus were not represented. This limits direct comparison of CTO mortality with that of elective PCI, which warrants further investigation. The database does not capture quality-of-life outcomes, such as functional status, or long-term results, precluding any cost-effective analysis of successful CTO intervention. Differences in these metrics between CTO and non-CTO patients remain an important area for future study. Finally, the NIS database lacks information on medical therapy, preventing assessment of potential treatment-related differences between groups.

## Conclusion

Analysis of a large national inpatient database with propensity score matching demonstrated that PCI for chronic total occlusion was associated with significantly higher mortality and postprocedural complications compared with PCI for non-CTO lesions. PCI should be considered only in carefully selected patients with severe, refractory angina unrelieved by optimal medical therapy.

## Data Availability

NIS database publicly available

---

**Figure:**
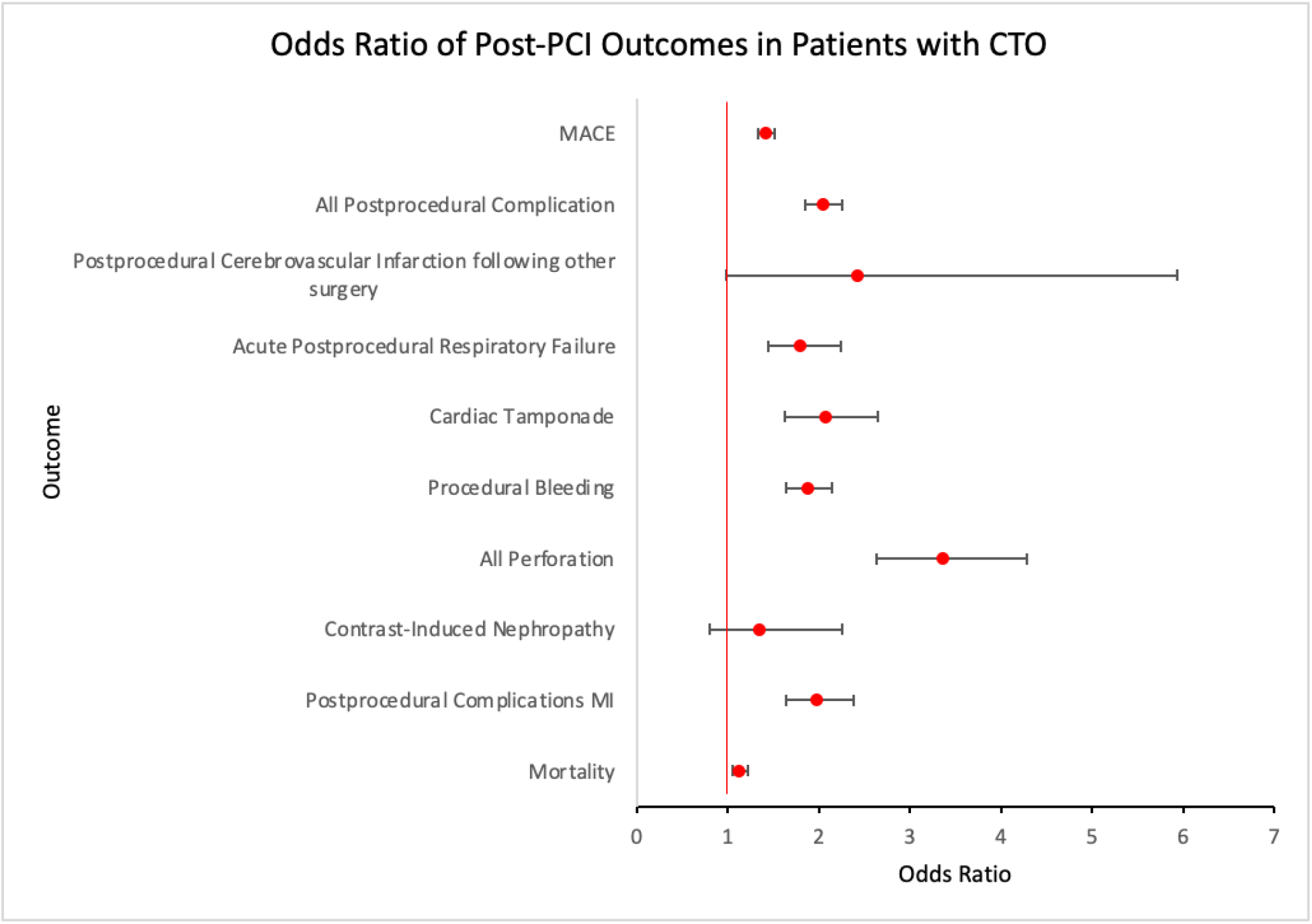
Figure showing odds ratios of outcome and complications data of patients undergoing percutaneous coronary intervention (PCI) of a chronically totally occluded vessel (CTO) with other non-CTO PCI

## References

1. Stone GW, Kandzari DE, Mehran R, et al. Percutaneous recanalization of chronically occluded coronary arteries: part I (consensus document). Circulation. 2005;112:2364–2372.

2. Namazi MH, Serati AR, Vakili H, et al. A Novel Risk Score in Predicting Failure or Success for Antegrade Approach to Percutaneous Coronary Intervention of Chronic Total Occlusion: Antegrade CTO Score. Int J Angiol. 2017;26(2):89–94.

3. Christofferson RD, Lehmann KG, Martin GV, Every N, Caldwell JH, Kapadia SR. Effect of chronic total coronary occlusion on treatment strategy. Am J Cardiol. 2005;95:1088–1091.

4. Ochiai M, Ashida K, Araki H, et al. The latest wire technique for chronic total occlusion. Ital Heart J. 2005;6:489–493.

5. Saito S. Different strategies of retrograde approach in coronary angioplasty for chronic total occlusion. Catheter Cardiovasc Interv. 2008;71:8–19.

6. Tsuchikane E, Suzuki T, Asakura Y, et al. Debulking of coronary chronic total occlusions with rotational or directional atherectomy before stenting: final results of the DOCTORS study. Int J Cardiol. 2008;125:397–403.

7. Jeroudi OM, Alomar ME, Michael TT, et al. Prevalence and management of coronary chronic total occlusions in a tertiary Veterans Affairs hospital. Catheter Cardiovasc Interv. 2014;84:637–643.

8. Grantham JA, Jones PG, Cannon L, Spertus JA. Quantifying early health status benefits of successful chronic total occlusion recanalization: the FACTOR trial. Circ Cardiovasc Qual Outcomes. 2010;3:284–290.

9. Sapontis J, Salisbury AC, Yeh RW, et al. Early procedural and health status outcomes after chronic total occlusion angioplasty: the OPEN-CTO Registry. JACC Cardiovasc Interv. 2017;10:1523–1534.

10. Christakopoulos GE, Christopoulos G, Carlino M, et al. Meta-analysis of clinical outcomes of patients who underwent percutaneous coronary interventions for chronic total occlusions. Am J Cardiol. 2015;115:1367–1375.

11. Godino C, Bassanelli G, Economou FI, et al. Predictors of cardiac death in patients with coronary chronic total occlusion not revascularized by PCI. Int J Cardiol. 2013;168:1402–1409.

12. Lee SW, Lee PH, Ahn JM, et al. Randomized trial evaluating PCI for chronic total occlusion: DECISION-CTO. Circulation. 2019;139:1674–1683.

13. Werner GS, Martin-Yuste V, Hildick-Smith D, et al. A randomized multicentre trial to compare revascularization with optimal medical therapy for chronic total coronary occlusions. Eur Heart J. 2018;39:2484–2493.

14. Rigger J, Hanratty CG, Walsh SJ. Common and uncommon CTO complications. Interv Cardiol. 2018;13:121–125.

15. Karacsonyi J, Vemmou E, Nikolakopoulos ID, Ungi I, Rangan BV, Brilakis ES. Complications of chronic total occlusion PCI. Neth Heart J. 2021;29:60–67.

16. Nathan A, Hashemzadeh M, Movahed MR. Percutaneous coronary intervention involving coronary bifurcation is associated with higher mortality and complications. Am J Cardiovasc Dis. 2024;14(3):180–187.

17. Khan MF, Brilakis ES, Wendel CS, Thai H. Comparison of procedural complications and in-hospital outcomes between successful and failed CTO-PCI: meta-analysis of observational studies. Catheter Cardiovasc Interv. 2015;85:781–794.

18. Nathan A Allistair,, et al. PCI of chronic total occlusion associated with higher inpatient mortality and complications compared with non-CTO lesions. Am J Med. 2023;136(10):994–999.

19. Lawton JS, Tamis-Holland JE, Bangalore S, et al. 2021 ACC/AHA/SCAI guideline for coronary artery revascularization. J Am Coll Cardiol. 2022;79:e21–e129.

20. Movahed MR. It is time to have better oversight and accountability in performing too many not indicated PCI in patients with CTO. Int J Cardiol. 2019;278:38–39.

21. Simsek B, Kostantinis S, Karacsonyi J, et al. A systematic review and meta-analysis of clinical outcomes of patients undergoing CTO-PCI. J Invasive Cardiol. 2022;34(11):E763–E775.

22. Henriques JPS, Hoebers LP, Råmunddal T, et al. EXPLORE: PCI for concurrent CTO in STEMI. J Am Coll Cardiol. 2016;68:1622–1632.

23. Mashayekhi K, Nührenberg TG, Toma A, et al. REVASC trial. JACC Cardiovasc Interv. 2018;11:1982–1991.

24. Al-Lamee R, Thompson D, Dehbi HM, et al.; ORBITA investigators. PCI in stable angina (ORBITA). Lancet. 2018;391(10115):31–40.

25. Xenogiannis I, Nikolakopoulos I, Krestyaninov O, et al. Impact of successful CTO-PCI on subsequent clinical outcomes. J Invasive Cardiol. 2020;32:433–439.

26. Juricic SA, Tesic MB, Galassi AR, et al. Randomized Controlled Comparison of Optimal Medical Therapy with Percutaneous Recanalization of Chronic Total Occlusion (COMETCTO). Int Heart J. 2021;62(1):16–22.

27. Park TK, Lee SH, Choi KH, et al. Late Survival Benefit of Percutaneous Coronary Intervention Compared With Medical Therapy in Patients With Coronary Chronic Total Occlusion: a 10-Year Follow-Up Study.

28. Movahed MR. Strong Bias Toward Performing Percutaneous Coronary Intervention in Patients With Chronic Total Occlusion Despite Lack of Important Benefit at a Very High Cost and Risk to the Patient. JACC Cardiovasc Interv. 2018;11(15):1540–1541.

29. Werner GS, Hildick-Smith D; Trial Investigators FTE. Reply: Due to the lack of significant mortality benefits along with high procedural complication rates, CTO-PCI should be discouraged. EuroIntervention. 2024;20(1):110.

30. Movahed MR. Excluding interventional harm in the West Denmark Heart Registry introduces major flaw. Catheter Cardiovasc Interv. 2025;106(1):286–287.

31. Movahed MR. Significant Downplay and Underreporting of Adverse Events in Patients Who Underwent Percutaneous Coronary Intervention of Chronic Total Occlusions. Am J Cardiol. 2024;210:317.

32. Movahed MR. Common practice of underreporting and downplaying adverse events and exaggerating benefits in patients undergoing percutaneous coronary intervention of chronic total occlusions. Scand Cardiovasc J. 2024;58(1):2373070.

33. Ellis SG, Ajluni S, Arnold AZ, et al. Increased coronary perforation in the new device era: incidence, classification (Ellis I–III/III-CS), management, and outcome. Circulation. 1994;90:2725–2730.

34. Kinnaird T, Anderson R, Ossei-Gerning N, et al. Legacy Effect of Coronary Perforation Complicating PCI for Chronic Total Occlusive Disease: Analysis of 26,807 Cases From the BCIS Database. Circ Cardiovasc Interv. 2017;10(5):e004642.

35. Kostantinis S, Simsek B, Karacsonyi J, et al. Incidence, Mechanisms, Treatment, and Outcomes of Coronary Artery Perforation During Chronic Total Occlusion Percutaneous Coronary Intervention. Am J Cardiol. 2022;182:17–24. doi:10.1016/j.amjcard.2022.07.004.

36. Movahed MR. Letter regarding “Chronic Total Occlusion Percutaneous Coronary Intervention: Present and Future”. Circ Cardiovasc Interv. 2025;18(7):e015552.

